# From Clinical Free Text to Auditable Concepts: An Agentic Framework for Interpretable Prediction

**DOI:** 10.64898/2026.09.02.26362086

**Authors:** Congning Ni, Weixin Liu, Qingyuan Song, Matthew Murrow, Bradley A. Malin, Zhijun Yin

## Abstract

Across application domains, predictive signals often sit in unstructured free text rather than structured fields, yet turning that text into useful and interpretable features is difficult. Running large language models (LLMs) over an entire corpus is costly and hard to reproduce, while end-to-end text representations can rely on surface cues that are difficult to inspect. We present an agentic workflow that takes a prediction task and a raw text corpus as input and produces an auditable feature layer. The first two agents use an LLM to derive a task-specific predictor taxonomy and weakly label a bounded text sample; routed local extractors then process the corpus, and a deterministic builder aggregates the evidence into a dynamic, longitudinal concept bottleneck. We evaluate the framework on medication discontinuation in a longitudinal oncology cohort and 30-day readmission in MIMIC-IV. With gradient boosting, the longitudinal bottleneck increases area under the receiver operating characteristic curve (AUROC) over coarse concept buckets from 0.700 to 0.761 for medication discontinuation and from 0.576 to 0.609 for readmission. The proposed framework achieves predictive performance comparable to direct BioClinicalBERT prediction on both tasks while additionally providing explicit, interpretable, and traceable task-specific concepts. LLM use is confined to a bounded weak-labeling stage costing $24.00 and $23.39, respectively, compared with projected costs of $10,648 and $11,519 for exhaustive sentence-level LLM processing of the full corpora, demonstrating the substantial cost efficiency of the proposed agentic system.

## 1 Introduction

For many prediction tasks, the signals needed for prediction are rarely confined to well-structured data fields. Large language models (LLMs) can read such text [19], but running them over a whole corpus is expensive, hard to interpret, and hard to reproduce [22]. This raises the question we study: *how can we convert a target task and a raw text collection into a representation that is task-specific, interpretable to the people who act on it, and auditable?*

We show that a single agentic pipeline can turn a prediction task and a raw text corpus into an interpretable feature layer, with no separate model hand-built for each new problem. The pipeline builds a *concept bottleneck* [7] of human-readable evidence from free text.

Building on agent-based decomposition [27] and LLM-driven agentic workflows [26], the pipeline runs as four specialized agents followed by a deterministic builder with no further LLM calls. A *Concept Taxonomy Agent (Agent 1)* proposes candidate concepts and extraction routes; a *Weak-Labeling Agent (Agent 2)* weakly labels a bounded snippet sample [15]; an *Extractor-Training Agent (Agent 3)* trains local models; and a *Corpus-Tagging Agent (Agent 4)* applies them across the corpus. The *Deterministic Builder* aggregates the tagged text into a structured concept layer. The resulting concept bottleneck is constructed from LLM-generated weak labels and extracted evidence rather than manually annotated per-instance concept labels.

We demonstrate the pipeline on early discontinuation of endocrine therapy in a longitudinal oncology cohort [11] and hospital readmission within 30 days of discharge in MIMIC-IV [5]. Because evidence builds up over time, the builder splits each broad concept into finer *sub-concepts* and aggregates them over preindex time windows [18]. This produces a dynamic, longitudinal representation whose contribution can be traced to individual concepts and source text.

This paper makes three contributions. First, we introduce a reusable agentic architecture that turns a target task and an unstructured corpus into an auditable concept layer through task-specific taxonomy construction, extraction routing, and bounded LLM weak labeling. Second, we provide a dynamic, longitudinal concept bottleneck that splits extracted evidence into sub-concepts and aggregates it over time without per-instance concept supervision. Third, we evaluate the framework on two clinical prediction tasks through prediction experiments, a direct-text BioClinicalBERT comparison, a sub-concept audit, and a quantitative analysis of computational cost.

## 2 Related Work

Agent-oriented design decomposes a complex task into specialized modules that exchange explicit intermediate artifacts [27]. LLM agents extend this idea by planning, calling tools, and coordinating sub-tasks [26], including in medical settings [30]. These systems generally solve tasks end-to-end; we instead use the same decomposition to construct an auditable feature representation for a separate downstream predictor. Named entity recognition tags predefined entity types at the token level [24], while pretrained encoders [3], LLM-based extraction [29], and weak supervision [15] broaden the schema or reduce labeling cost. Our pipeline instead derives the concept set from the task and routes each concept to a matched extraction unit.

Post-hoc methods such as LIME [16] and SHAP [9] explain a model after it is fit, but their explanations only approximate model behavior and can be unstable [17] or adversarially manipulated [20]. Concept bottleneck models instead route prediction through human-meaningful variables [7], while label-free variants remove the need to annotate those variables for every instance [12]. Recent work in vision similarly uses LLM-generated attributes and CLIP-derived concept scores, combined with residual representations, to improve robustness to spurious correlations [28]. However, these approaches construct static concept representations rather than a dynamic, longitudinal concept layer, in the sense of temporal abstraction [18], directly from raw text. Our builder addresses this gap in clinical prediction, where relevant signals often occur in narrative text and LLM-based processing remains difficult to audit at corpus scale.

Our two tasks instantiate this narrative setting but differ sharply in document structure. Early medication discontinuation, tied to toxicity and treatment burden [4] and previously predicted from structured data [11] or fine-tuned language models [10], is documented across many short notes per patient. Thirty-day readmission, tied to comorbidity [6] and discharge context [25], is concentrated in one or a few long discharge summaries per admission. These contrasting structures motivate an adaptive pipeline rather than a fixed schema.

## 3 Prediction Tasks and Clinical Cohorts

To evaluate the pipeline in a demanding setting, we apply it to two de-identified clinical prediction tasks (Table 1). The first predicts early medication discontinuation in an institutional oncology and specialty-medication cohort, defined as stopping treatment before completion of the planned course. The institutional review board reviewed this private-data study and approved it as non-human subjects research. The second predicts hospital readmission within 30 days of discharge using the publicly available de-identified MIMIC-IV cohort [5]. Each task defines an index date that anchors prediction: the treatment decision date for medication discontinuation and discharge from the index admission for readmission. The Corpus-Tagging Agent (Agent 4) processes the source note corpus for each task, after which the deterministic builder drops all post-index evidence before constructing the note-derived modeling representation.

**Table 1.**
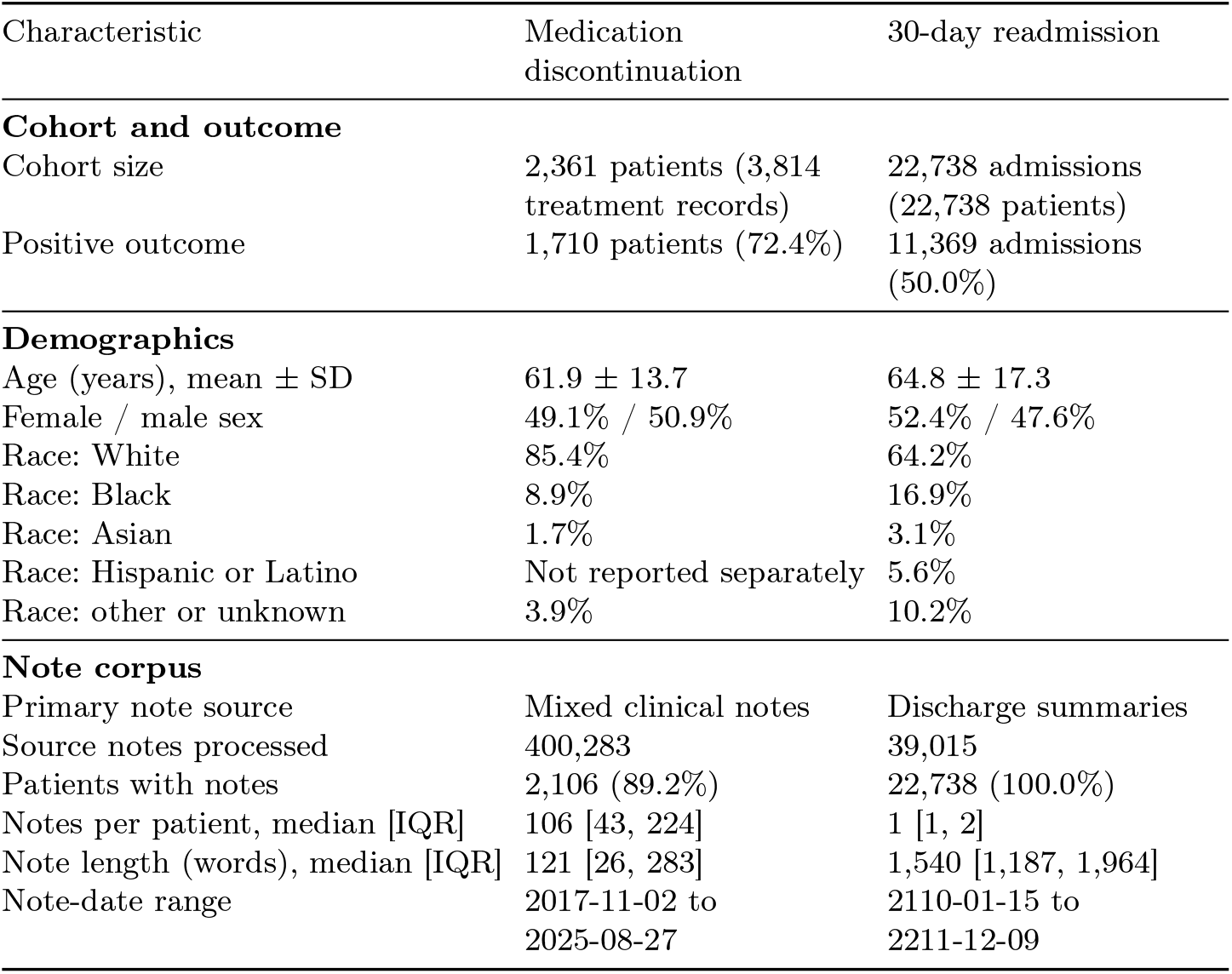
Cohort summarization. Medication-discontinuation positives are patients who discontinue treatment before completing the planned course. SD and IQR denote standard deviation and inter-quartile range, respectively. MIMIC-IV dates are de-identified and shifted into the future to preserve patient privacy.

The cohorts differ in both modeling unit and the distribution of textual evidence. Medication-discontinuation prediction is performed at the patient level, with evidence dispersed across many short longitudinal notes. Each MIMIC-IV admission instead forms one modeling row, with evidence concentrated in one or a few long discharge summaries. Applying the same representation framework to both settings therefore tests whether the dynamic-longitudinal bottleneck can accommodate diffuse longitudinal evidence as well as concentrated admissionlevel narratives.

## 4 Agentic Construction of the Note Concept Layer

Figure 1 summarizes the workflow. The *Concept Taxonomy Agent* and *Weak-Labeling Agent* use the LLM to define task-specific concepts and generate bounded weak supervision. The *Extractor-Training Agent* and *Corpus-Tagging Agent* then train and deploy local extractors across the source corpus. Finally, the *Deterministic Builder* assembles the extracted evidence into the dynamic-longitudinal bottleneck without further LLM calls.

**Fig 1.**
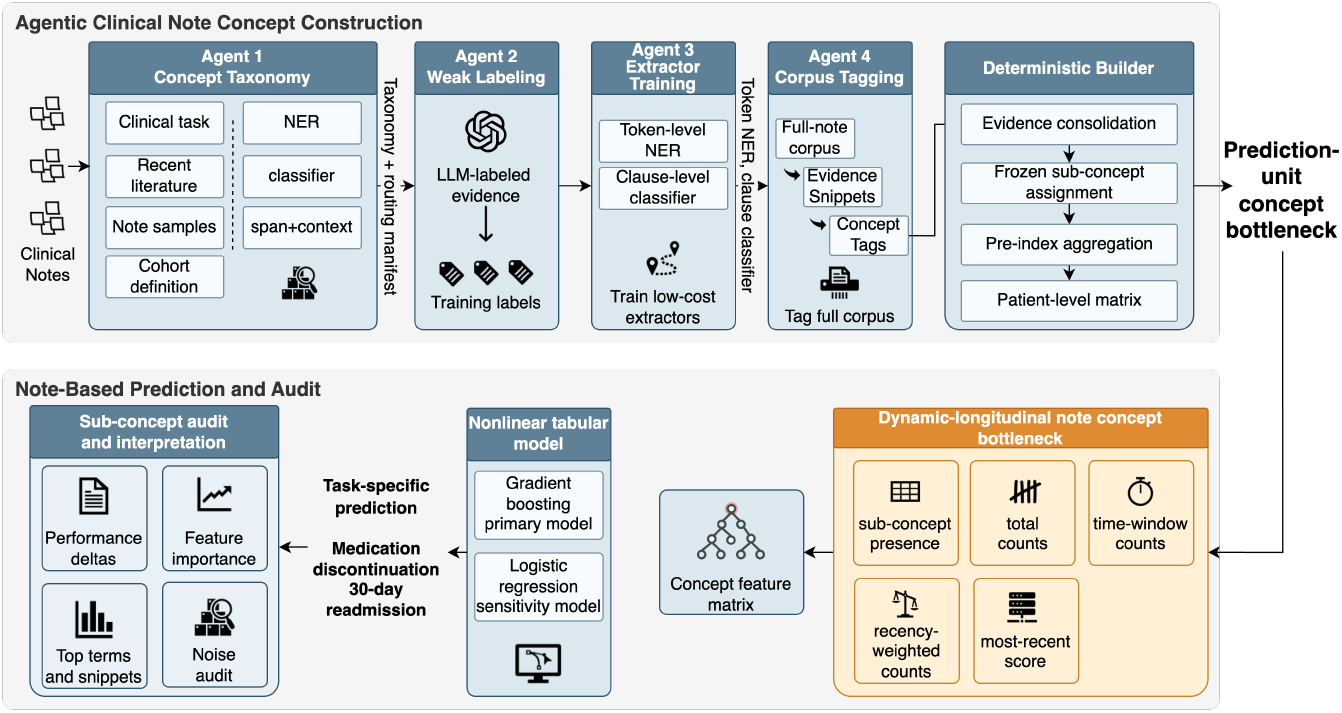
Overview of the four-agent pipeline and Deterministic Builder. The upper panel shows concept construction through corpus tagging; the lower panel shows the concept bottleneck, prediction model, and audit components.

### Concept Taxonomy Agent

The Concept Taxonomy Agent takes the prediction task, cohort definition, and a sample of cohort notes as input. It derives search terms from the task, retrieves recent PubMed abstracts, and uses the note sample to capture the cohort’s vocabulary and document structure. The agent uses GPT-5.3 to synthesize these inputs into a task-specific taxonomy and routing manifest. Local mentions are routed to token-level named entity recognition (NER), where negation and assertion remain local to the mention [2,24]; broader statements are routed to clause-level classifiers [23]; and hybrid concepts use both branches. Table 2 summarizes the resulting task-specific note schemas.

**Table 2.** Task-specific concepts produced by the Concept Taxonomy Agent.

| ID | Medication discontinuation concepts | 30-day readmission concepts |
| --- | --- | --- |
| $c_1$ | Prior adverse drug reaction or medication intolerance | Chronic medical conditions |
| $c_2$ | Prior cancer treatment exposure | Prior surgical or procedural history |
| $c_3$ | Baseline symptom burden | Medication use or regimen context |
| $c_4$ | Functional status or physical condition | Drug allergies or medication intolerance |
| $c_5$ | Chronic comorbidity burden in narrative context | Functional status and mobility |
| $c_6$ | Patient preferences or treatment hesitation | Substance use and behavioral health context |
| $c_7$ | Care coordination or access barriers | Social support and living context |
| $c_8$ | Healthcare utilization or acute clinical events | Care coordination and follow-up planning |

### Weak-Labeling Agent

The Weak-Labeling Agent generates weak training labels for the task-specific concepts. We sample note units from the training corpus and prompt the same LLM with the concept definitions, inclusion and exclusion rules, routing mode, and examples. The LLM returns concept-specific evidence labels with the corresponding short spans or clauses. This stage provides weak supervision for extractor training [15] rather than performing clinical inference. We bound LLM use by limiting the batch size and fraction of the corpus submitted, requiring a minimum number of examples per concept, and retaining the labeling logs as audit artifacts. Corpus-scale extraction therefore relies on local models rather than repeated LLM inference.

### Extractor-Training Agent

The Extractor-Training Agent trains the branches specified by the routing manifest by fine-tuning BioClinicalBERT [1], initialized from biomedical-domain pretraining [8], under the standard BERT framework [3]. The span branch uses beginning-inside-outside (BIO) token labels for local evidence, the segment branch classifies clause-level evidence, and hybrid concepts use both. Patient-disjoint splits reduce leakage risk; training includes entity-free negative units, early stopping, and validation-tuned confidence thresholds. A deployment manifest records the permitted branches for each concept, producing local extractors for corpus-scale tagging without further LLM inference.

### Corpus-Tagging Agent

The Corpus-Tagging Agent applies the trained extractors to the entire source note corpus and records the supporting snippet, patient and note identifiers, note date, concept, confidence score, and extraction branch. Before aggregation, low-confidence results, duplicates, repeated template text, and short spans without useful evidence are removed.

### Deterministic Builder

The Deterministic Builder drops all post-index evidence and converts the remaining note-level evidence into the modeling representation. Following fixed rules, it consolidates evidence across documents, organizes the evidence into sub-concepts, and computes count and recency features over the pre-index windows defined in the next section. It also produces the audit artifacts used downstream, including representative snippets, sub-concept prevalence, and top corpus terms.

## 5 Dynamic Longitudinal Bottleneck Design and Evaluation

Following the concept-bottleneck principle [7], the downstream model predicts *y*_*u*_*∈ {*0, 1*}* from 45 demographic covariates *a*_*u*_ and the pre-index concept representation *z*_*u*_, not raw notes. To isolate note-representation structure without structured electronic health record (EHR) event codes, we compare four nested conditions: demographics only; demographics plus 16 presence/count features for eight parent concepts; data-adaptive sub-concepts; and sub-concepts extended with recency and time-windowed features.

Broad parent concepts mix clinically distinct evidence, such as a stable home medication versus a newly started prescription. The builder decomposes each parent using mini-batch *k*-means over training-split snippet embeddings. From *K ∈ {*2, 3, 4, 6, 8, 12, 16 *}*, it selects the largest value for which every cluster contains at least 100 unique snippets and none exceeds 70% of the parent’s evidence; otherwise the concept yields no sub-concepts. Training-only centroids are frozen before validation and test assignment.

For longitudinal aggregation, the builder drops all post-index evidence and deterministically computes eight features per sub-concept: a presence indicator, a total count, a recency-weighted count, a most-recent score, and four time-windowed counts over the 0–30, 31–90, 91–180, and *>*180 day windows. A log(1+*·*) transform is applied to all counts. This procedure yields 110 sub-concepts and 880 total note features for medication discontinuation. For readmission, parent concepts *c*_4_ (drug allergies) and *c*_5_ (functional status) produced no extracted training evidence, so the resulting 96 sub-concepts and 768 total note features derive from the remaining six parent concepts.

The patient-disjoint training, validation, and test splits contain 1,239, 532, and 590 medication-discontinuation patients and 18,190, 2,274, and 2,274 readmission admissions; their positive rates are 72.2%, 72.6%, and 72.7%, and 50.0% in each split. The primary histogram gradient-boosting model uses 200 iterations, learning rate 0.04, and *L*_2_ regularization 0.05; class-balanced logistic regression is a linear sensitivity baseline. Models are fit on combined training and validation data and evaluated once on the test split. We report area under the receiver operating characteristic curve (AUROC) and area under the precision– recall curve (AUPRC), with 95% paired-bootstrap intervals and *p*-values from 1,000 shared-index test resamples without retraining.

As a direct-text comparison, BioClinicalBERT uses chronologically concatenated pre-index notes split into non-overlapping 512-token chunks, retaining up to eight recent chunks per patient. It is fine-tuned end to end on the combined training and validation splits for three epochs (learning rate 2 *×*10^*−*5^ and training batch size 16), with patient labels assigned to chunks; test chunk probabilities are averaged per patient. The same metrics and bootstrap procedure are used. For the direct-text comparison, BioClinicalBERT and the dynamic-longitudinal model are evaluated on the same text-eligible test subset for each task.

## 6 Results

### Prediction performance

Table 3 reports test performance under gradient boosting. Relative to parent concepts, the full dynamic-longitudinal representation improves AUROC by 0.061 for medication discontinuation (95% interval [+0.025, +0.095], *p*=0.002) and by 0.033 for readmission ([+0.010, +0.057], *p*=0.006). For medication discontinuation, most of the improvement comes from static sub-concepts (+0.043, *p*=0.010), while longitudinal features add a smaller nonsignificant increment (+0.018, *p*=0.236). For readmission, static sub-concepts contribute little (+0.009, *p*=0.364), whereas longitudinal aggregation provides the main gain (+0.025, *p*=0.004). Because the baseline includes demographics only, these results isolate the contribution of note-derived features rather than compare with a full structured-data model.

**Table 3.** Threshold-independent test performance under the primary gradient-boosted model. Each row cumulatively adds note features on top of demographics. Cells report point estimates and 95% paired-bootstrap intervals. AUPRC baselines are 0.727 for discontinuation and 0.500 for readmission.

| Representation | AUROC | AUPRC |
| --- | --- | --- |
| *Medication discontinuation* |  |  |
| Demographics | 0.635 [.583, .683] | 0.809 [.767, .851] |
| + Parent concepts | 0.700 [.656, .746] | 0.837 [.799, .877] |
| + Sub-concepts | 0.743 [.698, .790] | 0.864 [.826, .898] |
| + Sub-concepts + time | **0.761** [.720, .808] | **0.874** [.839, .909] |
| *30-day readmission* |  |  |
| Demographics | 0.541 [.517, .564] | 0.537 [.506, .568] |
| + Parent concepts | 0.576 [.553, .600] | 0.576 [.544, .609] |
| + Sub-concepts | 0.585 [.562, .608] | 0.587 [.555, .618] |
| + Sub-concepts + time | **0.609** [.588, .633] | **0.609** [.579, .645] |

Two additional comparisons provide context for the primary results. On the matched 485-patient medication subset with usable pre-index text, BioClinical-BERT scores 0.809 for AUROC and 0.896 for AUPRC, compared with 0.791 and 0.895 for the bottleneck; neither difference is significant (*p*=0.526 and 0.986). For readmission, the corresponding scores are 0.610 and 0.604 versus 0.609 and 0.609, again without significant differences (*p*=0.982 and 0.700). Direct text modeling therefore does not significantly improve predictive performance. In the linear sensitivity analysis, adding longitudinal features reduces medication-discontinuation AUROC relative to static sub-concepts (*−* 0.083, *p<*0.001) and is neutral for readmission (+0.001, *p*=0.944), suggesting that the longitudinal gain depends on interactions or sparsity patterns captured by gradient boosting.

### Sub-concept audit

We conduct a post hoc exploratory audit to examine which documentation patterns drive the gradient-boosted predictions. Sub-concepts are ranked by permutation importance on the held-out test split, measured as the AUROC decrease after permuting a sub-concept’s aggregation and window features. Because permutation importance is unsigned, test-set presence log odds ratios summarize association direction. Neither quantity is interpreted causally. For medication discontinuation (Fig. 2, upper), predictive contribution spans functional status, prior treatment, patient engagement, acute utilization, and symptoms; documented mobility is most important (*Δ*AUROC *≈*0.013). Functional status, engagement, and utilization appear less often in cases, whereas prior treatment has a positive but nonsignificant association. For readmission (lower), four of the top six sub-concepts concern medication-regimen context. Active medication changes (*p<*0.05) and chronic medication complexity (*p<*0.001) have unadjusted positive associations, indicating that distinctions within the coarse medication bucket matter more than medication presence alone. Representative snippets and terms also reveal repeated template text that can guide taxonomy or prompt refinement.

**Fig 2.**
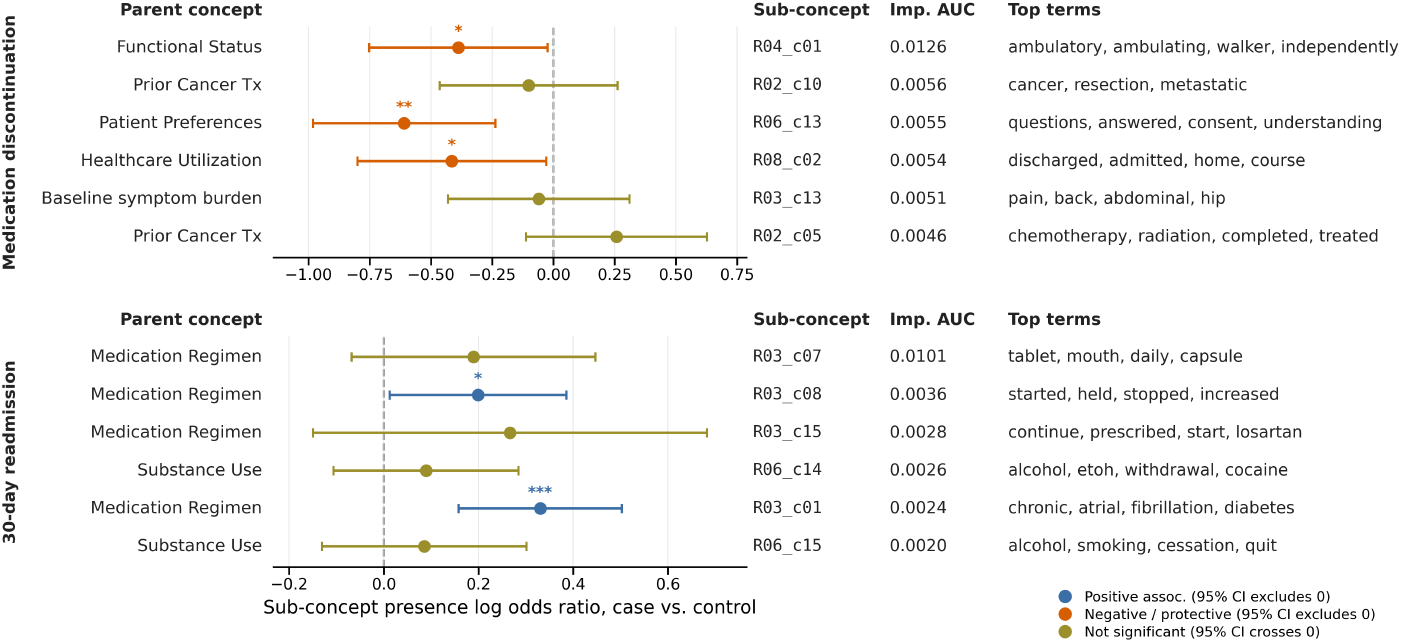
Post hoc test-set audit. Rows rank sub-concepts by permutation importance (Imp. AUROC); points show presence log odds ratios with 95% confidence intervals. Colors indicate unadjusted positive, negative, or nonsignificant associations (* *p <* 0.05, ** *p <* 0.01, *** *p <* 0.001); panels use independent scales.

### Computational cost

The bounded LLM stage remained small relative to corpus-scale processing. The Weak-Labeling Agent processed 9,112 of 10,000 medication-discontinuation candidate sentences for $24.00 and 8,591 of 10,000 readmission candidates for $23.39; both runs stopped when their fixed candidate pools contained no further eligible sentences. Training the two shared extractor branches (one NER model and one segment classifier, each covering all eight concepts) required 211.7 and 202.3 seconds, respectively. Without additional LLM calls, the Corpus-Tagging Agent processed 400,283 notes in 154.7 minutes and 39,015 discharge summaries in 92.1 minutes, producing 1,805,792 and 2,671,572 raw extractions. Assuming 15 words per sentence and applying the observed mean tokens per call and blended API prices, hypothetical exhaustive sentence-level LLM processing is projected to cost $10,648 and $11,519, corresponding to approximately 440 times the cost of our approach.

## 7 Discussion and Conclusion

Across two settings with different document profiles, the same construction process produced useful note representations. Medication discontinuation gained most from finer sub-concepts, while readmission gained more from temporal aggregation. This extends interval-based temporal abstraction [18] from structured measurements to extracted textual evidence.

The direct-text comparison clarifies the role of the additional representation-building steps. BioClinicalBERT did not significantly improve AUROC or AUPRC over the dynamic-longitudinal representation on either task, and the observed point estimates were close. The concept representation, however, keeps the intermediate evidence directly inspectable and supports the sub-concept audit. These comparisons do not establish equivalence between the two approaches, but they do not show a statistically significant predictive advantage for direct-text modeling in these experiments. The additional representation-building cost is limited through bounded LLM use. The language model weak-labels only a sampled candidate pool [21], following the weak-supervision setting in [15], while corpus-scale tagging is performed by local extractors. In our experiments, weak labeling cost $24.00 for medication discontinuation and $23.39 for readmission, whereas exhaustive sentence-level LLM processing was projected to cost $10,648 and $11,519, respectively, approximately 440 times as much. The local extractors then processed the full corpora without additional LLM calls. This separates the expensive LLM stage from corpus-scale extraction and makes repeated tagging more reproducible.

The second design choice is fixed-rule assembly. The concept layer is constructed before the downstream predictor is fit, so the features entering the model remain traceable to extracted evidence. The sub-concept audit uses this traceability to identify clinically coherent patterns, association directions, and documentation artifacts. A developer can inspect a low-importance or repeated-template cluster through its terms and representative snippets and then revise the taxonomy, routing manifest, or labeling prompts.

The agents construct the intermediate representation, while aggregation and prediction remain outside autonomous LLM decision making. Although the process requires only a task and document collection, this study evaluates it only clinically; non-clinical evaluation is needed before broader claims.

Several limitations remain. The demographics-only baseline isolates the contribution of note-derived features but does not measure their incremental value over a full structured-data model [13,14]. The extractors rely on weak rather than expert labels, and the stability of the generated taxonomy has not yet been systematically evaluated. Finally, the post hoc sub-concept audit uses the heldout test set and unadjusted associations and should therefore be interpreted as exploratory. Future work will evaluate multimodal models, taxonomy sensitivity, and concept extraction against expert annotations.

Despite these limitations, the main finding is consistent across both tasks. The agentic workflow converts free text into task-specific concept layers whose predictive value, temporal structure, and source evidence can be examined directly. The dynamic-longitudinal representation improves over coarse concept buckets, while direct BioClinicalBERT does not provide a statistically significant improvement over the concept representation. The framework therefore provides an auditable alternative to direct-text prediction while limiting LLM use to bounded weak labeling rather than full-corpus inference. Future work should add structured tabular baselines for multimodal prediction, evaluate the quality of extracted concepts with targeted human review, and test the workflow on non-clinical corpora.

## Data Availability

The institutional clinical data used for the medication-discontinuation analysis are not publicly available due to privacy and institutional restrictions. MIMIC-IV is available to credentialed users through PhysioNet subject to its data-use requirements.

## Acknowledgments

Omitted for double-blind review.

